# Fundamental limitations of case isolation

**DOI:** 10.1101/2021.03.31.21254677

**Authors:** Jonas Hansson, Alain Govaert, Richard Pates, Emma Tegling, Kristian Soltesz

**Affiliations:** Lund University, Dept. of Automatic Control, Lund, Sweden

**Keywords:** Fundamental limitations, network dynamics, case isolation, COVID-19

## Abstract

Case isolation is a strategy with the potential to curb infectious disease epidemics. Expressions for the stability boundary of a case isolation scheme defined through the proportion of the infectious population that it isolates with a given delay have recently been established. Here we quantify how this stability boundary moves when heterogeneity of the inter-individual contact network increases, and explain the underlying mechanism through insightful examples.

## 1. INTRODUCTION

### 1.1 Background

Early in an epidemic of a previously unknown disease, transmitted through a previously unknown pathogen, it can happen that existing pharmaceutical treatments are not applicable or efficient. This was the case with COVID-19, caused by the pathogen SARS-CoV-2. In early 2020 countries across the world experienced exponential increases in detected SARS-CoV cases, hospitalizations and ICU admittance due to COVID-19, and ultimately related deaths. While statistics are still debated one year later, as exemplified through Oliver (2021), it stood clear early on that pharmaceutical treatment and clinical care would not alone suffice to curb the epidemic outbreak.

The exponential increase of detected SARS-CoV-2 cases, and particularly of COVID-19 related deaths, during early 2020 resulted in governments implementing schemes to reduce the number of probable transmission events. Such schemes are within infectious disease epidemiology referred to as non-pharmaceutical interventions (NPIs for short), and can be partitioned into two categories:

- Recommendations or legislation aimed at decreasing inter-individual contact rates;
- Schemes for case isolation through testing, and possibly contact tracing.

The former includes, for example, discouraging unnecessary in-person interaction, banning large gatherings, closing schools or imposing societal lockdowns. The effectiveness of such interventions is still poorly understood. In particular, we have illustrated a lack of practical identifiability of individual NPI effectiveness from data obtained during the first European COVID-19 wave in Soltesz et al. (2020). This was the case despite a simplistic model formulation and optimistic assumptions on uncertainty in data.

In light of the above, case isolation schemes have gained attention. In contrast to NPIs aimed at reducing contact rates in general, the case isolation schemes target reduction of contacts involving infectious individuals, as schematically illustrated in Fig. 1. If successfully implemented, they could be the key to an open society where lockdowns are replaced by regular testing. Under such schemes, a positive test results in case isolation. The effectiveness of the scheme can then be further increased by complementing systematic testing with contact tracing.

**Fig. 1.**
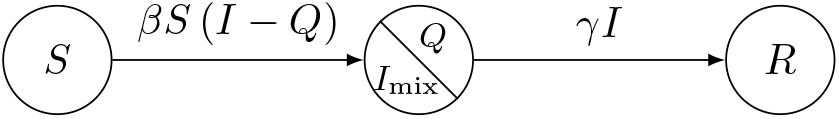
Schematic illustration of a case isolation scheme. The infectious proportion of the population *I* is partitioned into one isolated (quarantined) proportion *Q*, and one proportion *I*_mix_. The proportion *I*_mix_ interacts with the susceptible proportion *S*, and this interaction is governed by the mixing parameter *β*. The transition rate from the infectious *I* to the removed (recovered or deceased) proportion *R* is *γ*. We will focus on analysing how the design of the case isolation scheme that partitions *I* into *I*_mix_ and *Q* affects the evolution of the epidemic.

Notably, such a scheme has been demonstrated effective in, for example, New Zealand, as described by Robert (2020). At times when the case isolation scheme was deemed insufficient, New Zealand authorities instead enforced strict societal lockdowns.

The question we consider here is under what circumstances a case isolation scheme alone is sufficient to stabilize the epidemiological trajectory. We first derive criteria for an idealized case of a homogeneously interacting population, and then investigate how interaction heterogeneities affect these criteria.

### 1.2 Preliminaries

We consider a closed population. This is an optimistic approximation, since there will be no import cases within the model. The population is partitioned into susceptibles, infectious and removed (recovered and permanently immune or deceased) proportions. Here, we will not distinguish between being infected and being infectious. However, the results we are about to present can easily be extended to cater for this distinction.

If the proportion of infectious goes to zero, the strategy has been successful. The question we consider is how large the infectious proportion can grow, relative to the susceptible, in order for a case isolation strategy to remain effective.

To answer this, we consider the setting early in an epidemic, where the proportion of susceptibles is much larger than the infectious and removed, respectively. This allows us to disregard herd immunity effects. Another way to put it is that in an interaction between two individuals where one is infectious, the other is susceptible (with probability one).

Furthermore, we realistically assume the considered time window for our model to be short compared to the typical duration of immunity. This means that we neglect any flow from the removed to the susceptible sub-populations. As with distinguishing between infected and infectious individuals, introducing such reflux dynamics into the model we propose is straightforward, should the considered disease differ in this aspect from SARS-CoV-2.

We will focus on criteria for stabilization of the epidemic trajectory. We will refer to a case isolation scheme as stabilizing if it eventually empties the infectious subpopulation, without being aided by the herd immunity effect as introduced in Topley and Wilson (1923).

Note that this stability condition does not specify *performance* in the sense that it allows for an arbitrarily large infectious sub-population in the transient during which the infectious sub-population is emptied. As such, this stability condition constitutes a bare minimum. Any practically feasible strategy would need to fulfill it with some performance margin, as further elaborated in Pates et al. (2021).

Finally, we need to formalize our case isolation scheme. Here we will utilize a simple yet versatile model that quantifies the proportion of those infected on a given day that is subsequently isolated by the scheme *T*_delay_ days later.

This can be readily re-parameterized into, for example, the frequency with which individuals are tested, compliance to the scheme, logistic delays and specificity of the employed test for infection.

## 2. CASE ISOLATION IN HOMOGENEOUS POPULATIONS

### 2.1 Fundamental limitations

Assuming that the population is large, so that quantization effects become negligible, we can model the trajectory of the epidemiological state through

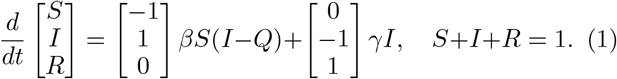

This is a slight variant of the traditional SIR model of Kermack and McKendrick (1927), where as usual *S, I* and *R* denote the proportions of the population that are susceptible, infectious and removed. The transitions between the states are governed by two rates that model the effect of disease spread and recovery, with mixing and recovery parameters *β* and *γ*, respectively. Note in particular that the mixing rate has been adjusted to account for the effect of isolating infectious individuals.

More specifically, *Q* denotes the proportion of the population that is both infectious and isolated, and the rate that describes the spread of the disease has been modified so that the spread is only driven by interactions between the remaining infectious population and the susceptible population (the *βS*(*I* − *Q*) term).

While simple, the formulation in (1) has a strength in that it allows various feedback strategies for regulating disease spread to be stated and analysed within a standard framework. Suppose, for example, that we wish to analyse the effectiveness of isolation schemes, in which infectious individuals are isolated as soon as they are identified as infectious. Arguably the simplest way to model this is through the feedback law

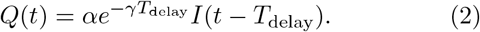

In words this equation states that the isolated population at time *t* is equal to some proportion 0 ≤ *α* ≤1 of the infectious population *T*_delay_ days in the past.^1^ This captures to a first approximation the two most important features of a case isolation scheme, namely, how quickly infectious individuals are identified and isolated (*T*_delay_), and what proportion of cases are found (*α*).

It is important to note here that cases in this context is equivalent to infectious individuals. This should not be confused with *detected* cases. For example, a detection rate of 80 % imposes an upper bound of *α* = 0.8. Particularly, *α* = 1 could only be achieved in a population where *all* cases are detected and *every* individual complies to the scheme.

By analysing (1)–(2) together we can begin to build our intuition for how the isolation scheme parameters *α* and *T*_delay_ affect disease spread for a disease with parameters *β* and *γ*. For example (as is well known) if no isolation occurs (*α* = 0), then at the start of an epidemic the infectious population will initially grow exponentially according to

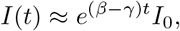

where *I*_0_ is the size of the initial outbreak. A natural question is then how large *α* must be (or how short *T*_delay_) before the period of exponential growth can be averted. This was the question studied in (Pates et al., 2021, §2.1.2), where it was shown that the equations (1)– (2) are locally asymptotically stable in a population in which *S ≫ I* if and only if

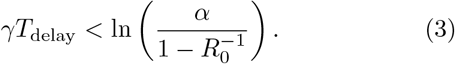

In the above *R*_0_ = *β/γ* is the basic reproduction number of the disease in the absence of isolation measures.

The specific trade-off between parameters and delay implied by (3) is shown in Fig. 2. This figure can be used to quickly assess the amount of delay that can be tolerated before instability, and hence exponential growth, occurs.

**Fig. 2.**
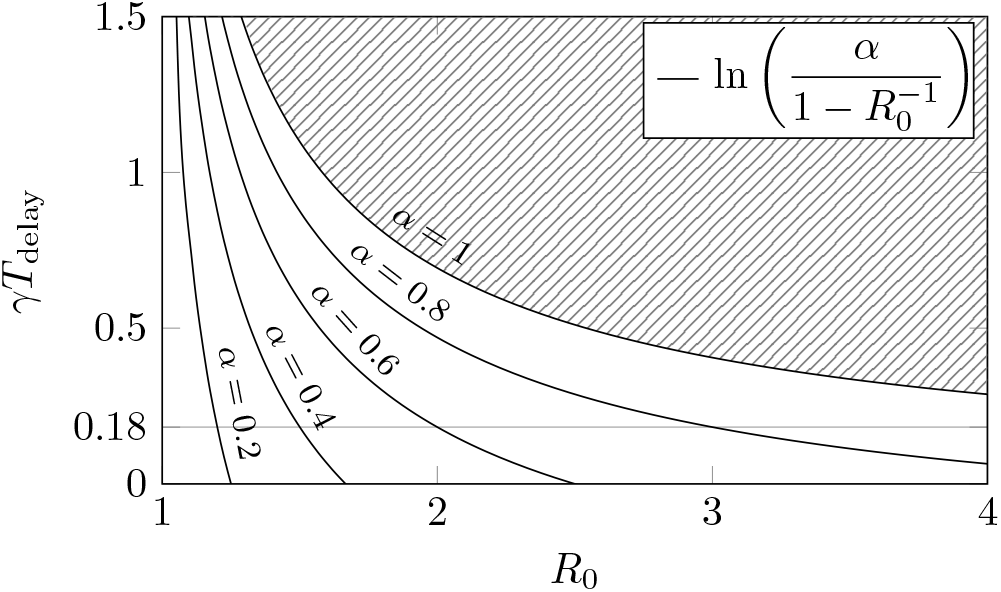
Illustration of the stability boundary for the model in (1)–(2). The model is stable if and only if (*R*_0_, *γT*_delay_) lies below the corresponding *α* curve. That is, at least a proportion *α* of persons becoming infectious on a particular day need to be isolated *T*_delay_ days later, given a natural recovery rate is *γ* days^*−*1^.

For example, with *α* = 0.8, *R*_0_ = 3 and *γ* = 0.1, parameters chosen to be representative for SARS-CoV-2, the stability condition becomes

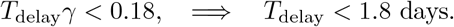

Clearly, short isolation times are essential when dealing with an infectious disease! We also see the importance of identifying a significant proportion of cases. By the time *α* ≈ 0.7 (that is, the scheme detects and isolates 70 % of the cases) exponential growth will occur even with *T*_delay_ = 0.

### 2.2 Notes on the reproduction number

The reproduction number is a common source to confusion, and therefore deserves explicit attention: In general, the reproduction number *R* describes the expected number of secondary infections caused by one primary infection. It holds that

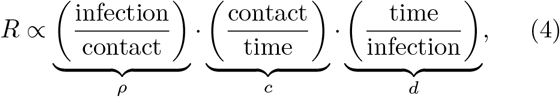

where

- *ρ* ∈ [0, 1]: probability of transmission given contact between a susceptible and infectous individual;
- *c* ∈ ℝ_+_: average contact rate between susceptible and infectious individuals;
- *d* ∈ ℝ_+_: average duration of infectiousness.

For the homogeneous population model (1) we have that *β* = *ρc* and *d* = 1*/γ*. It is also evident from (4) that the reproduction number reveals how *much* an epidemic grows, and not how *fast*. In order to quantify the latter, the serial interval between infections is needed in addition. Yet, the reproduction number is more commonly used than the corresponding growth rate *r* = *β* − *γ* among infectious disease epidemiologists, which is why we have chosen to use the former in our parameterization of fundamental limitations.

The basic reproduction number *R*_0_ is the reproduction in absence of a *considered* intervention. There is a common misconception that for a particular pathogen *R*_0_ is a universal constant (that can be looked up in the literature). Instead it depends on, among other time-varying parameters, the virulence of the pathogen and the societal structure under consideration. For instance, a particular virus would typically result in different *R*_0_ in a two countries, or in a city versus a village.

If an intervention is enacted, it is instead common to talk about the resulting effective reproduction number *R*_*e*_ (sometimes referred to as the time-varying reproduction number *R*_*t*_), and it really only makes sense to consider *R*_0_ in relation to a particular intervention: *R* = *R*_0_ in absence of the intervention; *R* = *R*_*e*_ if the intervention is enacted.

It was shown in Pates et al. (2021) that for the case isolation scheme (2) applied to the model (1) the relation between *R*_0_ and *R*_*e*_ is given by

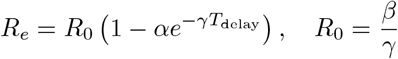

which is less than 1 if and only if (3) holds.

## 3. CASE ISOLATION IN HETEROGENEOUS POPULATIONS

### 3.1 Fundamental limitations

The model introduced in the previous section assumes a homogeneously interacting population. This is clearly a simplification. In reality, the interactions that may lead to infection are much more complex and hard to model. The probability to interact with someone at your workplace is for example much higher than to interact with someone from a remote country. There is also a time-varying aspect. For instance, if your workplace issues a work-from-home guideline to reduce disease transmission, your probability to interact with someone from your workplace will typically drop.

Numerous works have been dedicated to modelling the associated contact network dynamics, with May (2006); Salathé et al. (2010) and survey Nowzari et al. (2016) constituting representative examples. The validity of such network models is hard to verify, and the time-varying aspects further increases the uncertainty surrounding their accuracy. We therefore delimit ourselves to a simple but important question: how does introduction of interaction heterogeneity alter the requirements on our case isolation scheme (2), expressed in terms of the isolation proportion parameter *α* and associated time delay *T*_delay_?

To account for contact rate heterogeneity, early models of infectious diseases (particularly STDs) in May and Anderson (1987); May et al. (2001) incorporated a distribution of contact rates *p*_*k*_ = *N*_*k*_*/N* being the proportion of the population of size *N*, who on average have *k* contacts per time unit and in all other regards are homogeneous. As in Anderson et al. (1986) the corresponding disease dynamics of the homogeneous case generalize to

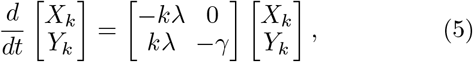

where *X*_*k*_ and *Y*_*k*_ denote the number of susceptible and infectious individuals in partitions *k* = 1, 2, …, *n*, respectively. The parameter *λ* = *β ∑*_*k*_ *kY*_*k*_ / *∑*_*k*_ *kN*_*k*_ is the probability of an infection acquired from any one randomly-chosen contact—now more likely to come from the partition with higher contact rates.

Early on in the epidemic it holds *X*_*k*_ = *N*_*k*_, which allows the collective epidemic dynamics to be written as

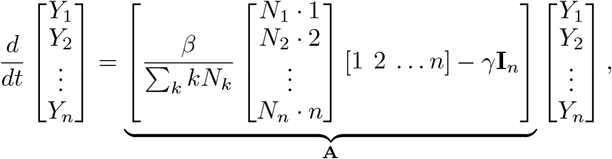

where **I**_*n*_ is the (*n* × *n*) identity matrix. The eigenvalues of **A** now determine the trajectory of the epidemic early on. They are given by −*γ* and

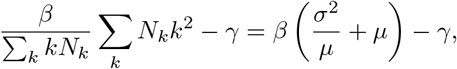

where *µ* = ∑_*k*_*kp*_*k*_ and *σ*^2^ = ∑_k_*k*^2^*p*_*k*_ −*µ*^2^ denote the mean and variance of the contact rates respectively. It follows that the linearised system about the disease-free equilibrium (*X*_*k*_ = *N*_*k*_ for all *k*) is stable if and only if

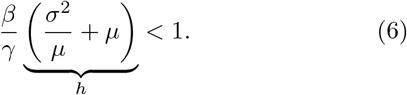

As originally described in May and Anderson (1987) and May et al. (2001), the adjustment factor *h* ≥ 1 in (6) thus increased the mixing parameter *β* = *ρc* of (4) relative to the homogenous case, where {*c* = *µ, σ* = 0} ⇒ *h* = 1. Since *R ∝ ρc* = *β*, an equivalent interpretation is that the reproduction number is increased by *h*.

The adjustment by the factor *h* remains valid under the case isolation scheme (2), since the probability to isolate an infectious individual is independent of the contact degree of that individual. Consequently, the stability condition of Theorem 2.1 in Pates et al. (2021) becomes

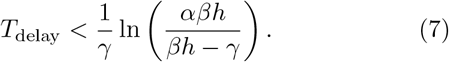

The derivative of the upper bound on the delay with respect to the coefficient of variation, or relative standard deviation, *c* _*υ*_ = *σ/µ* is

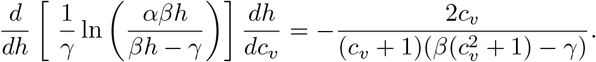

This implies that heterogeneity (*c* _*υ*_ > 0) *increases* the upper bound on *T*_delay_ if and only if the reproduction number of the corresponding homogeneous system (*c* = *µ*) in absence of control (2) fulfils *R*_0_ *<* 1*/h*. Since *c*_*c*_ *υ >* 0 ⇔*h >* 1 this requirement implies that heterogeneity allows for a longer *T*_delay_ in the control law (2) only if the uncontrolled system is already stable in the sense that *R*_0_ *<* 1. On the other hand, and of larger practical importance, the upper bound on admissible *T*_delay_ *decreases* when heterogeneity (*c*_*v*_) is increased whenever *R*_0_ > 1.

In the latter case, one may ask how large the coefficient of variation can be to allow for a positive delay and a stable equilibrium (*I, R, Q*) = (0, 0, 0) of the linearised model with the scaled parameter *β*. That is,

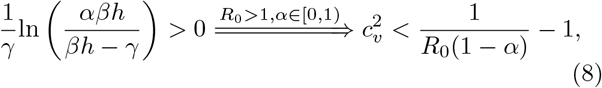

where, again, *R*_0_ is the basic reproduction number for a corresponding homogeneously interacting population.

Fig. 3 illustrates this maximum allowed coefficient of variance *c*_*v*_ of the contact graph, as a function of the basic reproduction number and proportion of cases *α* that are isolated according to (2).

**Fig. 3.**
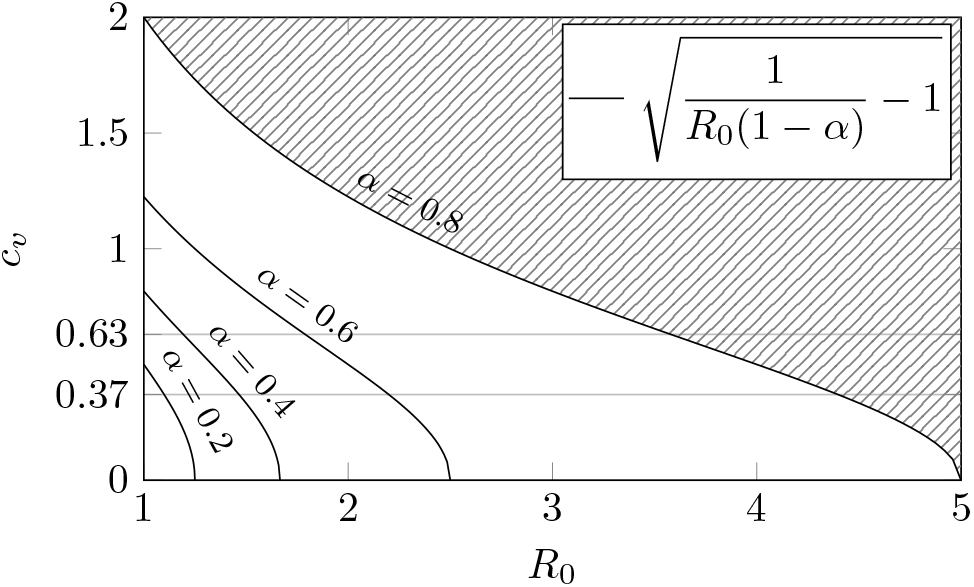
Illustration of maximum admissible coefficient of variance *c*_*v*_ for stabilization using the case isolation scheme (2) early in an epidemic (when *S ≫ I*). In this example, we assume that a proportion *α* of the infectious population are isolated without delay (*T*_delay_ = 0). The figure illustrates, for different *α*, the relation between the basic reproduction number *R*_0_ of a corresponding homogeneously interacting population (in absence of case isolation), and the admissible *c*_*v*_. The shaded curve shows what combinations of *R*_0_ and *c*_*v*_ cannot be stabilized for *α* = 0.8. The horizontal lines at *c*_*v*_ = 0.37 and *c*_*v*_ = 0.67 represent bounds on *c*_*v*_ for human infectious disease transmission networks reported in Salathé et al. (2010).

Revisiting the example from Sec. 2 with *γ* = 0.1 and *R*_0_ = 3, the condition (8) reveals that the maximum coefficient of variation for which a positive isolation delay *T*_delay_ > 0 is allowed evaluates as *c*_*v*_ ≈0.82. As a reference, Salathé et al. (2010) reports lower and upper bounds of 0.37 and 0.63 for *c*_*v*_ in human infectious disease transmission contact networks. The corresponding lines have been added to Fig. 3 and indicate that if *R*_0_ = 3 was computed or estimated for a homogeneous population, delay-free (*T*_delay_ = 0) isolation of a proportion somewhere in the range 0.6 *< α <* 0.8 would be required to stabilize the epidemic under (2).

### 3.2 Notes on contact heterogeneity

The phenomenon of the effective mixing parameter *β* increasing with increased heterogeneity (quantified by the coefficient of variation *c*_*v*_) is also the answer to the question “Why your friends have more friends than you do”, explained with mathematical insight in Feld (1991). The intuition is that individuals with many contacts are more likely to get infected, and therefore the infectious proportion of the population will comprise individuals who have more social contacts (higher degree) than those in the susceptible proportion of the population.

As with friendships, the contacts considered here are mutual, translating into an undirected network graph. Thus, the early spread of the disease is proportional to the mean number of contacts of a randomly chosen contact of an infectious individual (node), given by *h* in (6). Treating the network as homogeneous (*c* _*v*_ = 0 ⇔ *h* = 1) can therefore lead to an under-estimation of the reproduction number by disregarding the growth of the infectious subpopulation, which is fueled by highly connected individuals that are both more likely to acquire and spread infection.

In Fig. 4 we have illustrated this effect through simulations of the early stage of 100 epidemics on an undirected random graph representing a population. The nodes of the graph represent individuals; the edges contacts.

**Fig. 4.**
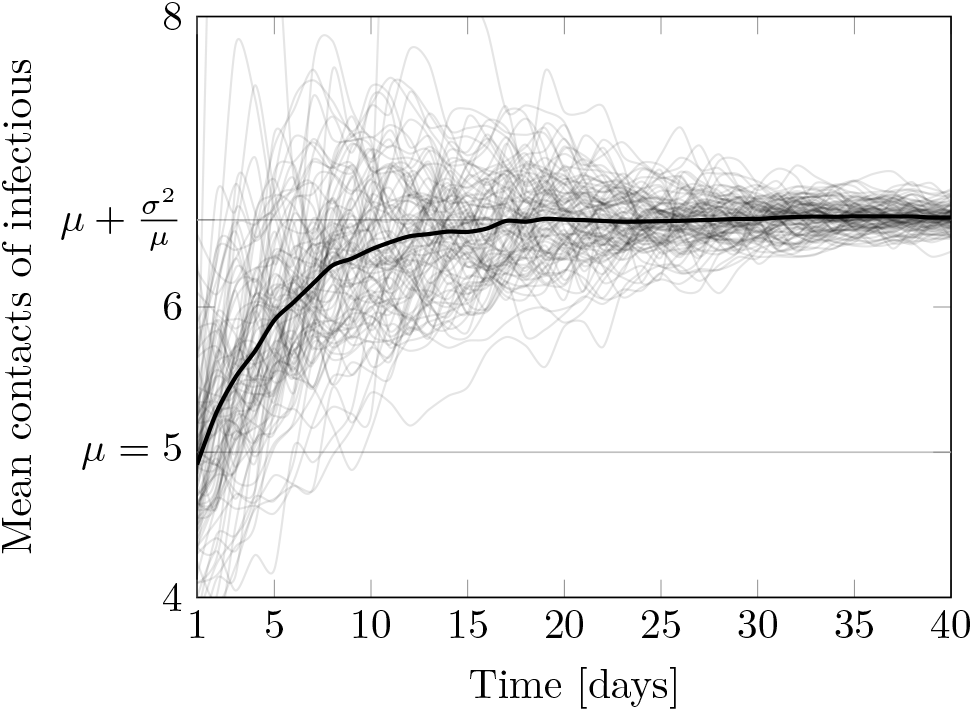
Simulation of 100 epidemics (grey) over an Erdős Rényi graph where the mean number of connections to the infectious (black) are shown each day. At the beginning of the epidemic this amounts to the mean contact degree *µ* of the graph, but soon stabilizes at the expectatation *µ* + *σ*^2^*/µ* of the excess degree distribution of the network, with *σ*^2^ being the contact degree variance of the network.

For the example in Fig. 4, we chose to generate Erdős Rényi graphs of *n* = 2 .10^6^ nodes and *M* = 5 .10^6^ undirected edges due to their simplicity, but other types of graph generation methods could also be used to illustrate the phenomenon.

The epidemics were each seeded by randomly assigning 10 infectious individuals on day one. This corresponds to *I* = 5 .10^*−*6^ ≪ *S* = 1 − *I*. The transmission probability from (4), being the per-day probability that an edge between one infectious and one susceptible node leads to disease transmission, was set to *ρ* = 0.05. The recovery rate from (1) was set to *γ* = 0.1. Assuming independent interactions, the probability for a susceptible to become infectious the next day, given that it has *m* infectious contacts, is thus 1 − (1 − *ρ*)^*m*^.

The cumulative infectious proportion was retrospectively evaluated to satisfy *I* + *R* 0.01 for each of the 100 simulations, validating the assumption of a fully susceptible population (or early epidemic stage) on which the stability analysis has been based.

In Fig. 4 the results from 100 epidemics simulations are shown. In particular, we see the average number of contacts of the infectious population throughout the early stages of the epidemic. From this figure we can clearly identify how the infectious individuals have a higher degree of contacts than the general population of the network. Furthermore, we can see that average number of contacts of the infectious population tends towards the expected value of the excess degree distribution, being the distribution of edges from a node reached by following an edge.

## 4. DISCUSSION

In Sec. 2 we characterized the stability boundary (3) associated with the case isolation scheme (2) as a function of the basic reproduction number *R*_0_ in absence of case isolation, and a recovery rate *γ* defined through (1).

In Sec. 3 we then characterized how the stability boundary is moved if *R*_0_ of a homogeneous population is used in (3), when in fact the population is heterogeneous in the sense that the coefficient of variation of the contact network *c*_*v*_ is positive, while the mean degree remains unchanged.

Introducing heterogeneity in this way corresponds to multiplication of the mixing parameter *β* in (1) with the scaling factor 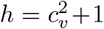. Since *β* is directly proportional to *R*_0_, the heterogeneity can be interpreted as increasing the reproduction number by the corresponding factor. Note that the same shifting applies for a population partitioned based on heterogeneity of the infectiousness parameter *ρ*. This makes it possible to apply the same modelling to account for a variability in infectiousness across variants of the considered pathogen.

Whether to multiply by the factor *h* before using *R*_0_ in the analysis comes down to how *R*_0_ was estimated from data. If it was estimated taking the heterogeneity into account it should not be adjusted, otherwise it should.

In the heterogeneous model (5) proposed in Anderson et al. (1986), the interaction between individuals with many and few contacts respectively are stochastic, and therefore do not correspond to a fixed (time-invariant) contact graph. For the case of a fixed contact graph, the stability bound (7) obtained for (5) would be conservative because of the assumption that the disease can spread to any contact of an infectious individual. In particular, an infectious individual cannot re-infect its own infector. Therefore, when considering epidemics on configurator model graphs, the shift of the epidemic threshold needs to be corrected by −1, resulting in 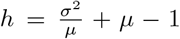, as further discussed in Newman (2018); Meyers (2007); Kiss et al. (2017).

Relatedly, clustering of the contact network result in the bound (7) becoming conservative. When many neighbours in the network are shared, (e.g. as in small-world networks) local clustering coefficients in the contact network are high and voids the assumption that all neighbors of an infectious node are susceptible. To analytically quantify how local and global clustering effects affect the stability bound of the studies case isolation schema along the lines of Trapman (2007) requires more complex network models. It falls outside the scope of this contribution, but constitutes a worthwhile direction for future work.

## 5. CONCLUSION

While obtaining detailed graph models of (the time-varying) human contact graphs relevant for infectious disease transmissions is generally not tractable, control theoretic analyses can provide qualitative, and to some extent quantitative criteria for the feasibility of strategies aimed at halting disease spread. This has been illustrated here by expressing stability conditions for case isolation schemes in homogeneous and heterogeneous populations, as functions of fundamental epidemiological parameters.

## Data Availability

N/A

## 6. ACKNOWLEDGMENTS

This work was partially funded by the Swedish Research Council (grants 2017-04989, 2019-00691) and by the Wallenberg AI, Autonomous Systems and Software Program (WASP) funded by the Knut and Alice Wallenberg Foundation. The authors are members of the ELLIIT Strategic Research Area at Lund University.

## Footnotes

1 The 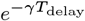 term is correcting for the fact that some of the infectious population will have recovered over these *T*_delay_ days (recall that *Q* is the sub-population that is both infectious and isolated).

